# Magnetic Resonance Imaging Simulation of the Skull Base, Oral Cavity, Pharynx, and Larynx: Advanced Imaging Protocol for a Radiation Oncology Center

**DOI:** 10.64898/2026.10.05.26364781

**Authors:** Natalie A. West, Lucas McCullum, Johnny T. Cheruvelil, Vi T. Dinh, Yao Ding, Jihong Wang, Komal Shah, Kim Learned, Melissa Chen, Jack Phan, Anna Lee, Adam Garden, David Rosenthal, Clifton D. Fuller

**Author notes:** contributed equally (Natalie A. West, Lucas McCullum).

## Abstract

Head and neck radiation oncology, with its complex locality and epidemiology, requires various imaging techniques to achieve sufficient visualization. Through collaboration with medical physicists, radiation therapists, radiation oncologists, and neuroradiologists, our group clinically implemented a standard set of magnetic resonance imaging (MRI) sequences on an MR-simulation scanner for three anatomical regions: skull base, oral cavity, and larynx/pharynx. A total of 41 patients with head and neck cancer and 3 volunteers were scanned on either a 1.5T or 3T MR-simulation scanner. Sequences described include Dixon-based approaches, SPAIR-based approaches, short T2-based approaches, and quantitative imaging methods. We describe and outline the steps to optimize these notable MRI sequences to aid in successful implementation in a radiation oncology clinic.

## Introduction

The head and neck region is a complex locality that requires numerous MRI-based contrasts to visualize its many anatomies^1^. This is further complicated by the epidemiology of head and neck cancers, including squamous cell carcinomas of the oral cavity, larynx, pharynx in addition to skull base tumors^1^. This becomes exceptionally important in radiation therapy (RT) simulation for head and neck cancers when differentiating gross target volume from normal tissue. MRI is especially equipped for this due to its superior soft tissue visualization compared to traditional computed tomography (CT)-based simulation. This enhanced visualization allows for distinct margins and sparing to healthy tissues. Additionally, MRI can reconstruct multiple sequences within a single acquisition, achieving various contrasts, reducing the number of imaging exams and multi-modality registration errors.

Recent advances in RT, including stereotactic radiosurgery (SRS), stereotactic body RT (SBRT), image-guided RT (IGRT), and particle (i.e., proton) therapy have improved target coverage and normal tissue sparing, effectively shifting treatment for tumors away from surgery and toward RT^2^. This, along with the inability of CT-simulation to effectively visualize soft tissue, has resulted in an unmet need for an MR-simulation program for RT treatment planning. We aim to develop three standard protocols for the radiation oncology clinic, including 1) skull base protocol, 2) oral cavity, and 3) larynx and pharynx, on an MR-simulation device to be used for RT treatment planning and post-RT normal tissue toxicities.

## Methods

Where possible, all acquisitions were prioritized to be isotropic reconstructed voxel size to view suprahyoid pathology and naso-ethmoidal neoplasms with equal resolution. The notable exceptions were the diffusion-weighted imaging (DWI) acquisition due to time constraints and signal-to-noise-ratio (SNR) concerns, the multi-dynamic multi-echo (MDME) acquisition due to low SNR, black bone mandible due to very high in-plane resolution, and dynamic contrast-enhanced (DCE) MRI due to time constraints. The abbreviated sequences are shown in **Tables 1–3** with a selection of sequence parameters for the skull base included in the **Supplementary Materials**.

**Table 1.**
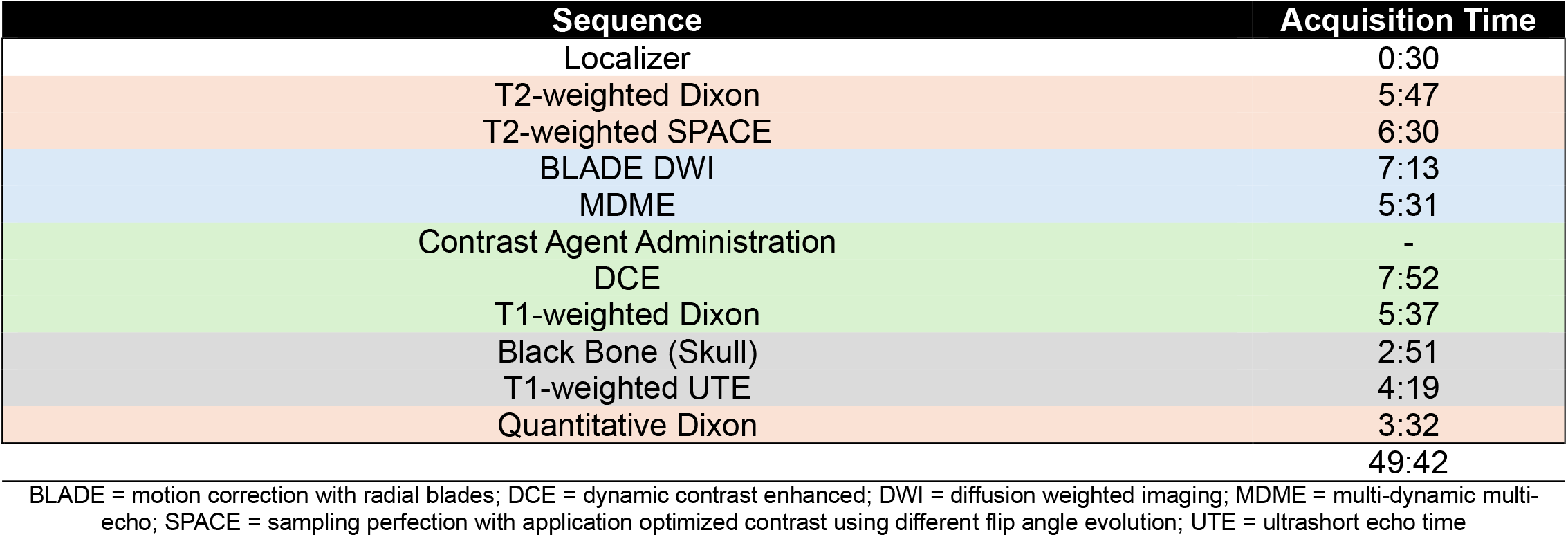
A listing of the MRI sequences used in the skull base protocol as well as their respective acquisition times. The sequences are divided into orange cells for sequences with fat suppression, blue cells for functional / quantitative imaging, grey cells for sequences used for bone evaluation and alignment with CT, and green cells for post-contrast sequences. Note that the T1-weighted Dixon is both a fat suppressed and contrast-enhanced sequence, though it is labeled here as only contrast-enhanced.

**Table 2.**
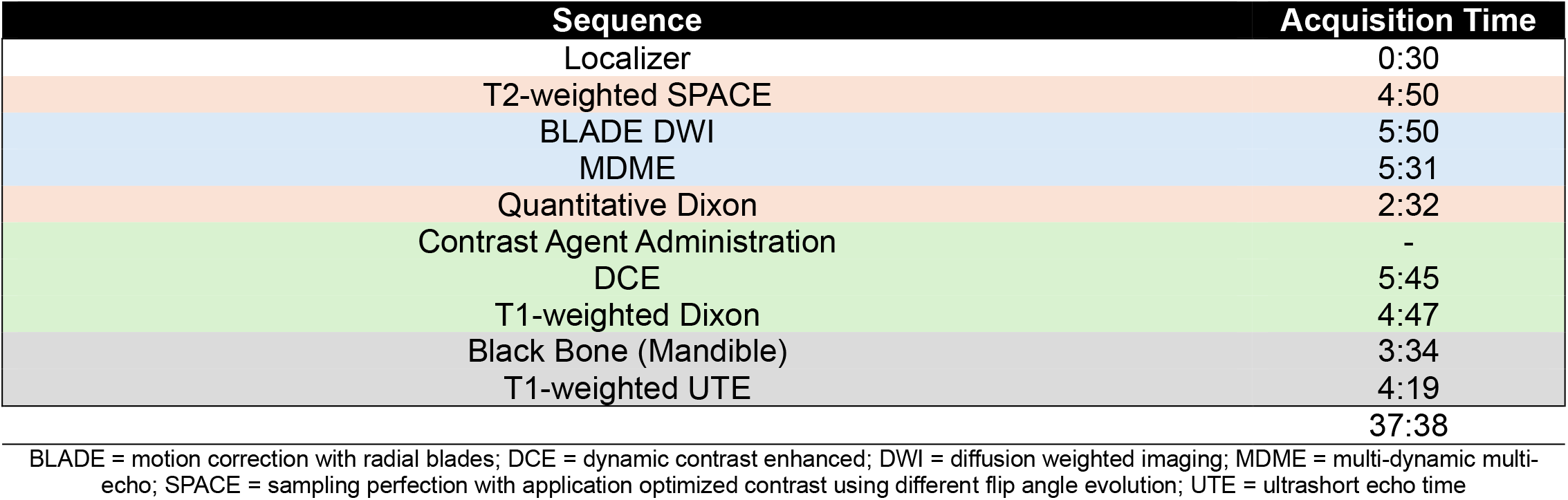
A listing of the MRI sequences used in the oral cavity protocol as well as their respective acquisition times. The sequences are divided into orange cells for sequences with fat suppression, blue cells for functional / quantitative imaging, grey cells for sequences used for bone evaluation and alignment with CT, and green cells for post-contrast sequences. Note that the T1-weighted Dixon is both a fat suppressed and contrast-enhanced sequence, though it is labeled here as only contrast-enhanced.

**Table 3.**
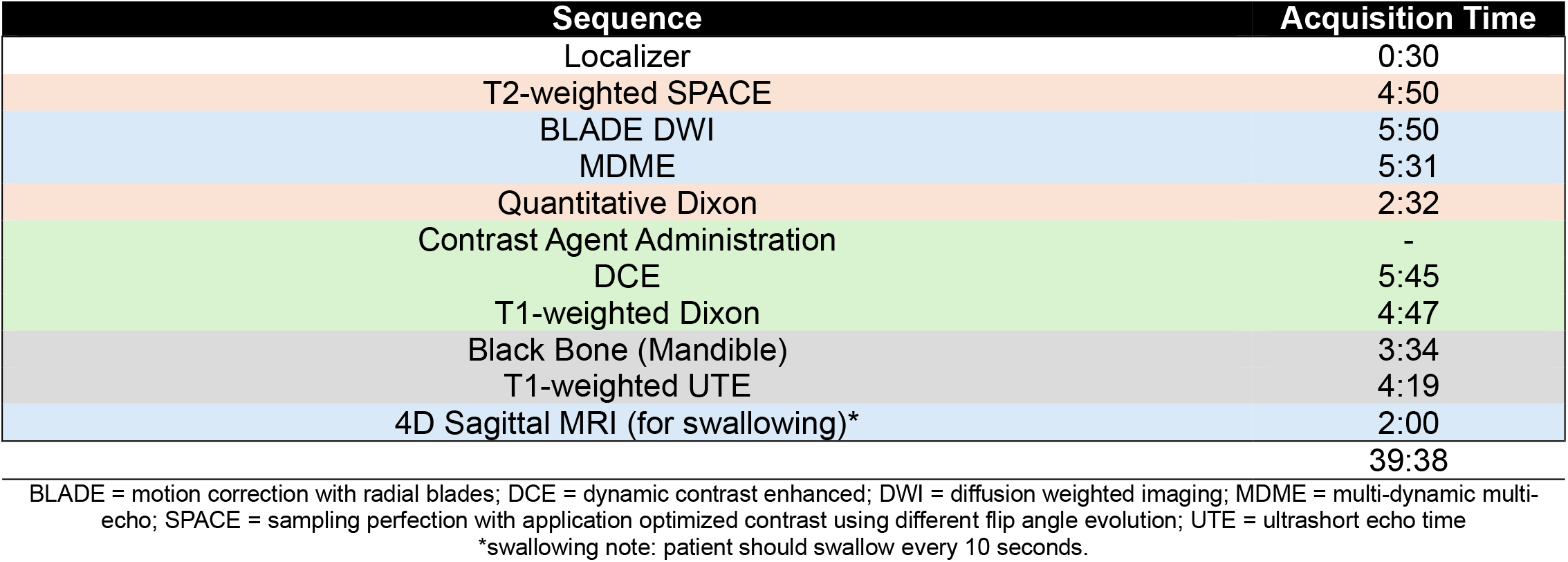
A listing of the MRI sequences used in the larynx/pharynx protocol as well as their respective acquisition times. The sequences are divided into orange cells for sequences with fat suppression, blue cells for functional / quantitative imaging, grey cells for sequences used for bone evaluation and alignment with CT, and green cells for post-contrast sequences. Note that the T1-weighted Dixon is both a fat suppressed and contrast-enhanced sequence, though it is labeled here as only contrast-enhanced.

This study followed protocols approved by the University of Texas MD Anderson Cancer Center Institutional Review Board for volunteers (PA15-0418) and patients (2024-0406). All participants were prospectively enrolled and provided written informed consent.

To demonstrate clinical feasibility, 41 patients with head and neck cancer were scanned on a 1.5T MR-simulation scanner (MAGNETOM Aera; Siemens Healthineers, Erlangan, Germany) or a 3T MR-simulation scanner (MAGNETOM Vida; Siemens Healthineers, Erlangen, Germany). Three volunteers were scanned on a 3T MR-simulation scanner (MAGNETOM Vida; Siemens Healthineers, Erlangen, Germany) and one volunteer had three repeat scans. Most examinations were performed using two UltraFlex Small 18-channel coils positioned on the bilateral cheeks. However, for larger subjects, an additional UltraFlex Large 18-channel coil was added to cover the shoulder and upper chest region. Volunteer scans were qualitatively reviewed by three radiation oncologists and one MRI physicist for clinical acceptability for MR-simulation-specific use cases. Prior work by our group on clinical feasibility and repeatability and reproducibility assessments, notably with MDME^3,4^, black bone^5^, and BLADE DWI^6^, was used to support clinical integration.

## Results

Fat suppression in head and neck cancer is a useful, and arguably necessary, technique which is often applied to increase contrast between tumors and surrounding normal tissue, particularly for tumors in the parapharyngeal space or bone marrow^7^. This is due to most head and neck tumors exhibiting intermediate high T2 and high T1 signal intensity similar to fat signal intensity in the normal surrounding tissue. Further, the head and neck extracranial space offers an abundance of fat-water interfaces which define the anatomical contrast in the region. Additional contrast while using fat suppression can further elucidate tumors in the perineural space^8^.

There are many methods of fat suppression in MRI, including T1-based methods which attempt to nullify the signal from fat by applying an inversion recovery pre-pulse and beginning the excitation pulse when the fat signal crosses zero. However, this method fails when the fat tissue T1 values are inhomogeneous since only a single inversion time can be chosen. Luckily, fat and water resonate at slightly different precession frequencies resulting in a chemical shift which can be exploited to better nullify fat using the so-called spectral approaches. In this section, we describe the sequences that we have chosen with added fat suppression using two techniques and how each of these are intended to be used to benefit the head and neck MRI-simulation workflow.

### 1. Dixon-Based Approaches

One of the most commonly used and foundational methods of fat suppression in MRI is using the so-called “Dixon”-based approach originally proposed in 1984 by Thomas Dixon^9^. Typical Dixon images are acquired with fat and water resonant frequencies both in-phase and out-of-phase, also known as opposed-phase^9^. This method, known as the “2-point” Dixon since it acquires two separate images at different resonance offsets between water and fat, estimates the water and fat only images from algebraically combining the in-phase and opposed-phase images. The water-only image is calculated by the summation of in-phase and opposed-phase while the fat-only image is calculated by subtracting the opposed-phase image from the in-phase image.

Moving beyond theoretical assumptions, clinically applying this technique exhibits relative insensitivity to both B0 and B1+ inhomogeneities^10^; however, its scan time may be longer due to the additional in-phase and opposed-phase information, thus increasing TR. One limitation of the traditional 2-point Dixon method is that the fat spectrum is complex and composed of multiple resonances peaks which cannot be sufficiently captured using only two images. Additionally, T2* can significantly influence the resulting fat fractions measurements and should be corrected for^11^. Therefore, modern advanced methods use a 6-point method^12^ to accurately capture all of the relevant fat peaks. Clinically, the use of this technique has been demonstrated in phantoms and healthy human subjects to be sufficiently repeatable and reproducible as well as correlating significantly to other confounders such as age in some structures^13^.

**Figure 1.**
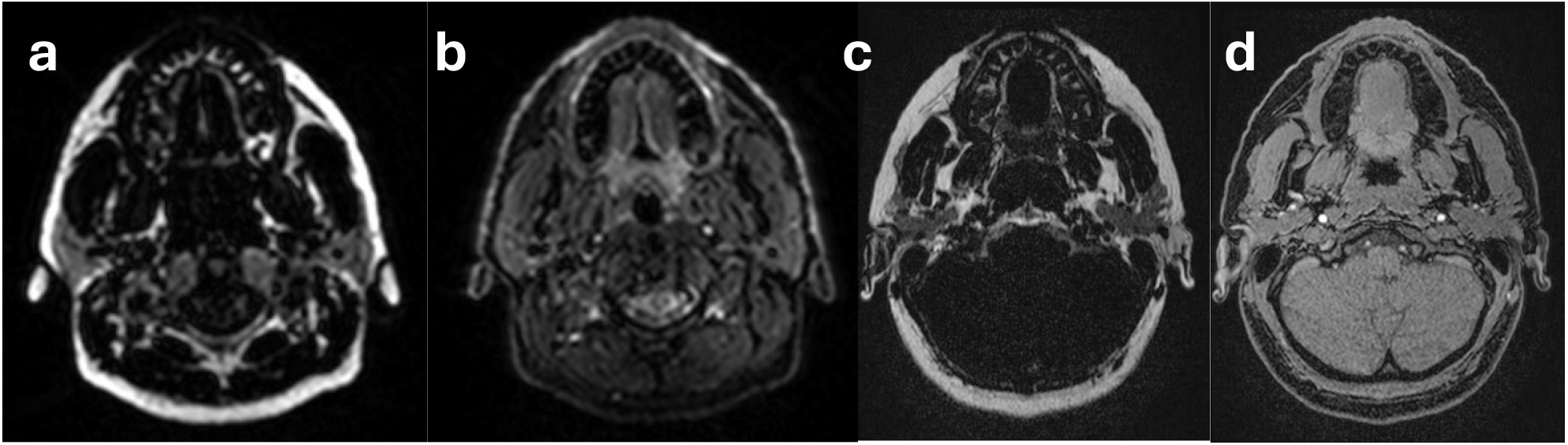
An example of fat (a) and water (b) reconstructions derived from the T2-weighted Dixon and fat (c) and water (d) reconstructions derived from the T1-weighted Dixon in the head and neck region.

#### a. Pre-Contrast / T2-weighted

From existing literature in the pre-contrast space, T2-weighted Dixon provides the optimal contrast for general evaluation in the head and neck region. A study utilizing a three-point T2-weighted Dixon technique demonstrated improved fat suppression and lesion conspicuity for the spine, neck, and orbit regions^14^. Another study showed that, even with a two-point Dixon method, vertebral lesions can be differentiated from normal vertebrae using the fat fraction^15^. These findings have been corroborated by another which added that the T2-weighted Dixon images can even replace conventional T1-weighted spin echo images for this purpose^16^.

#### b. Post-Contrast / T1-weighted

The use of gadolinium-based contrast enhancement can be used to characterize intracranial extension which can be better visualized in some cases^17^. In our case, our physicians plan to use this sequence for special use cases, such as to evaluate post-surgical patients. The preference for T1-weighted MRI following the administration of gadolinium-based contrast agents instead of T2-weighted MRI is due to how these contrast agents affect MRI signal properties, mainly by shortening the T1 relaxation time more profoundly than T2.

Combining gadolinium-based contrast agents with the Dixon fat suppression technique has been shown to exhibit more uniform fat suppression in the submandibular and supraclavicular regions in axial and coronal T1-weighted MRI images compared to spectral-based approaches^10^.

### 2. SPAIR-Based Approaches

Another method of fat suppression, known as STIR (Short T1 Inversion Recovery), can be achieved by applying an inversion pre-pulse with a time (TI) calculated as: TI = ln(2) * T1fat where T1fat is ~300 ms at 1.5T and ~360 ms at 3T^18^. Taking this a step further, SPectral Attenuated Inversion Recovery (“SPAIR” or “SPIR”) is another method of fat suppression which utilizes spectrally selective adiabatic inversion pulses instead of conventional inversion pulses^19^. This addition helps to offset the effects of B1+ inhomogeneities, resulting in more homogeneous fat suppression in larger volumes, at the expense of a higher specific absorption rate (SAR) in the patient. The benefit of the spectrally selective inversion pulse is that the maximum amount of water signal remains for maximum signal-to-noise ratio (SNR).

#### a. Conventional T2-weighted

In the head and neck, this technique with a T2-weighting has been used to identify osteoradionecrosis (ORN) often presenting as mixed signal intensities in the mandible^20^, for its diagnostic ability to detect metastatic lymph nodes^21^, and for evaluating extension of nasopharyngeal carcinoma (NPC) into the clivus and pharyngobasilar fascia^22^. An additional secondary application of this sequence to the head and neck region is the visualization of the brachial plexus and lumbosacral plexus given a sufficient field-of-view^23^.

#### b. T2-weighted SPACE

Optimal isotropic voxel sizes can be achieved by utilizing the “Sampling Perfection with Application optimized Contrast using different flip angle Evolution (SPACE)” method on Siemens systems, “CUBE” on GE, and “Volume Isotropic Turbo spin echo Acquisition (VISTA)” on Philips which utilizes a fast spin echo (FSE) contrast mechanism with a slab selective variable excitation pulse, turbo factors exceeding 100, and a low specific absorption rate (SAR). To make isotropic 3D MRI imaging faster using FSE, long echo trains should be utilized to make the best use of the short echo spacing. However, one major complication with this approach is that longer echo trains introduces blurring artifacts due to the low available signal remaining in the later echoes. To counteract this effect, variable flip angles based on the known T2 decay properties of the tissues being imaged can be applied to help maintain constant signal across the echo train, thus preserving T2 contrast at higher echo train lengths, enabling faster isotropic imaging^24^. This method additionally reduces SAR due to the lower flip angles being used instead of the typical, near 180°, pulses for higher SNR. One unfortunate side effect of modulating the flip angles is that the FSE sequence becomes sensitive to motion effects, particularly in the cerebral spinal fluid (CSF) in the midbrain and cervical spine causing the typically bright CSF to become dark^24^.

Despite this drawback, several studies have found this technique to be superior in expert evaluated visualization of the cranial nerves^25^. In head and neck cancers, a possible source of spread is through the nerves^26^, therefore, visualizing the nerves accurately helps to assess the degree of invasion into nearby structures. Cranial nerves around the skull base are surrounded by CSF, allowing them to be easily shown using a T2-weighted contrast which helps to separate the nerve from the CSF. In our protocol, we decided to include both the T2-weighted SPAIR and the T2-weighted SPAIR-SPACE to have both the heavy T2-weighting from the non-SPACE sequence with the compromise of image blurring, as well as the clearer isotropic voxels from the SPACE sequence with the compromise of CSF motion sensitivity to best delineate the cisternal segments of the cranial nerves.

### 3. Bone Injury Evaluation

Bones in the head and neck can be sites of injury, such as osteoradionecrosis, or disease, such as skull base tumors. While current conventional MRI methods cannot adequately visualize bone, recent advances in short echo time (TE) and low flip angle imaging techniques have been developed to overcome this challenge.

Notably, black bone^27^, and ultrashort echo time (UTE)^28^ MRI show promise as accurate, high-resolution methods in bone visualization. When successful, the implementation of these techniques allows for bone disease and injury monitoring and aid in CT-MRI registration via bony alignment.

#### a. Black Bone

Black bone MRI features a low flip angle, short repetition time (TR), and short TE and has been shown to adequately visualize cortical bone comparable to CT^5,27^. We implemented black bone MRI at a 0.33x0.33x1.00mm^3^ resolution, allowing for routine monitoring of small changes in cortical bone indicative of osteoradionecrosis^5^. Additionally, we extended the field-of-view (FOV) of black bone MRI to include the orbits and clavicle at a 1 mm isotropic resolution to aid in skull base treatment planning.

#### b. T1-Weighted UTE

Short echo time imaging techniques, including 2D/3D Ultrashort Echo Time (UTE), Zero Echo Time (ZTE), and Pointwise Encoding Time Reduction with Radial Acquisition (PETRA), have been similarly developed for bone morphometric and quantitative functional assessment^29^. Due to its TE of under 0.1ms, UTE captures bone, with its short T2 values, before it decays. Our group has implemented a sagittal T1-weighted UTE MRI for both mandibular and skull base visualization, as its radial acquisition allows for a large FOV.

**Figure 2.**
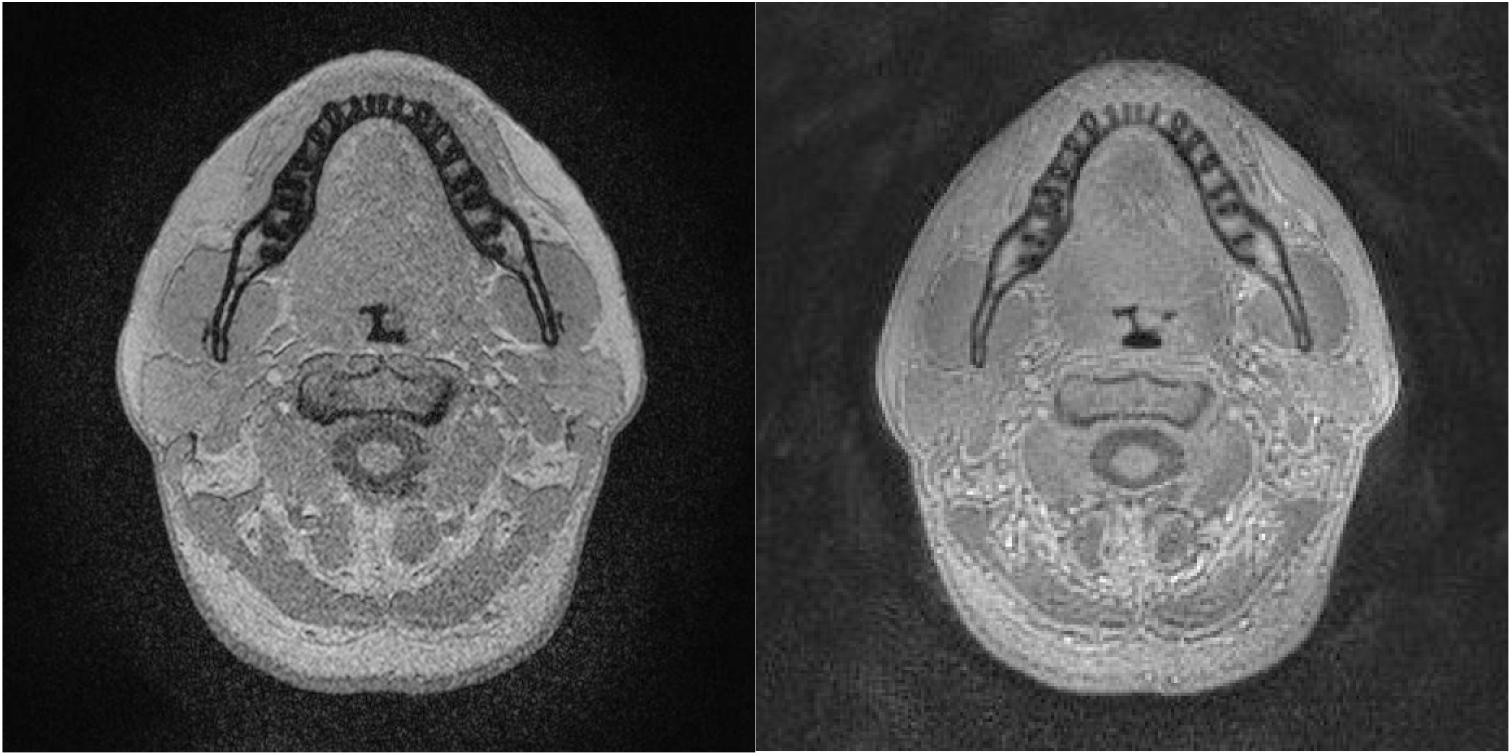
Black bone (left) and UTE (right) MRI for mandibular cortical bone visualization.

### 4. Quantitative Imaging

The applications of MRI in head and neck cancer imaging are traditionally qualitative, relying heavily on contrast-weighted images. However, qualitative MRI is not repeatable or measurable due to it having arbitrary units. Therefore, recent work has been directed towards increased utilization of more quantitative techniques for head and neck cancer evaluation^30^. Repeated quantitative measurements can subsequently be used as biomarkers which can be applied to measure changes in tumor size, function, and response to therapies^25^.

This framework can be applied to enhance analysis of clinical trials, especially those including MRI-guided radiation therapy to monitor not only tumor effects but also normal tissue changes like lymphedema and fibrosis^31^. Additionally, pairing quantitative maps with anatomical images can be used to help identify where the tumor is and enhance its contouring accuracy. For these reasons, we have included three quantitative MRI acquisition methodologies as part of our institution’s MRI head and neck cancer simulation protocol.

#### a. Diffusion Weighted Imaging (DWI)

DWI is an established MRI technique routinely used in the radiation oncology space. Despite its low resolution, DWI enables high contrast, particularly for tumor visualization and treatment response. DWI is a quantitative functional MRI technique that images the diffusion of primarily water molecules across membranes. Through using multiple b-values, DWI produces an apparent diffusion coefficient (ADC) map representing tissue diffusion. Our institution previously used an echo planar imaging (EPI) based approach (Readout Segmentation of Long Variable Echo-Trains, or RESOLVE) for ADC mapping which resulted in severe geometric distortion that was unable to be resolved through acquisition parameter optimization. The breaking point for this sequence for our institution was when it was used to contour a head and neck tumor and subsequently resulted in a near-miss event upon treatment plan review due to the significant level of distortion compared to the CT simulation scan. Therefore, during protocol optimization, we decided to switch to the motion correction with radial BLADEs (BLADE) technique which uses a non-Cartesian radial k-space acquisition strategy to minimize geometric distortion. However, significant subsequent optimization had to be done to achieve a similar scan time and SNR as the previous RESOLVE technique due to its slow acquisition speed.

**Figure 3.**
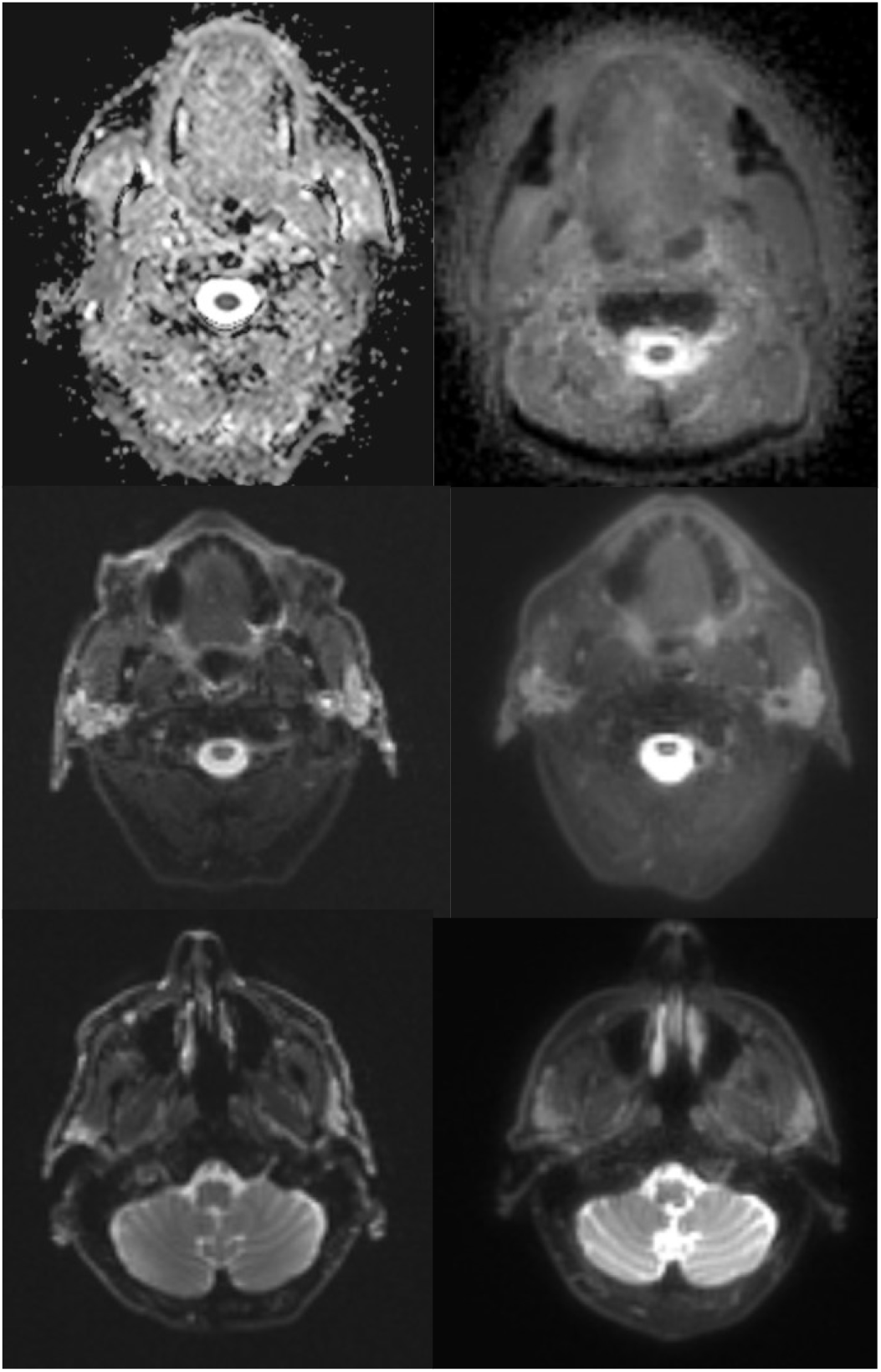
Previous EPI-based method (left) compared to updated BLADE-based method (right) at three different slice locations.

#### b. SyntheticMR: T1, T2, PD

SyntheticMR is an MRI technique developed by SyntheticMR AB (Linköping, Sweden) which simultaneously generates quantitative T1, T2, and PD maps^32^. Fundamental tissue properties can be measured using quantitative MRI and have been correlated with water content, macromolecular / lipid presence, and temperature^33,34^. An important side note is that T1 increases with the static magnetic field strength while T2 stays relatively constant within clinical ranges up to near 7T^35^. From these quantitative maps, their post-processing software, SyMRI, can generate synthetic contrast-weighted images which are interactable to the user through their user interface. Users can retrospectively adjust the TR and TE of the images to evaluate different contrasts as well as introduce one or two inversion pulses with customizable inversion times to nullify desired tissues. This MRI acquisition technique is commonly known as “MDME” on Siemens systems, “MAGiC” on GE, and “SyntAc” on Philips.

This technique and its synthetic contrast-weighted images have been verified in a prospective, multi-center, multi-reader trial^36^. Further, recent studies have validated the technical feasibility of this technique^3^ and its optimal acquisition parameters^4^ across the radiation oncology department, specifically in phantoms and for the head and neck. In addition to its feasibility at the phantom level, multi-participant studies have assessed the technique in head and neck cancers^37^. Supplementary applications of this technique have shown it to elucidate radiation-dependent demyelination in normal appearing white matter in glioma patients^38^ as well as its ability to assess the performance of automatic contouring algorithms across different contrast-weighted MRI images^39^. One recent study from Wei et al. showed that combining the quantitative maps from SyntheticMR with the ADC maps from DWI can enhance predictive value of discriminating metastatic and non-metastatic lymph nodes^40^.

**Figure 4.**
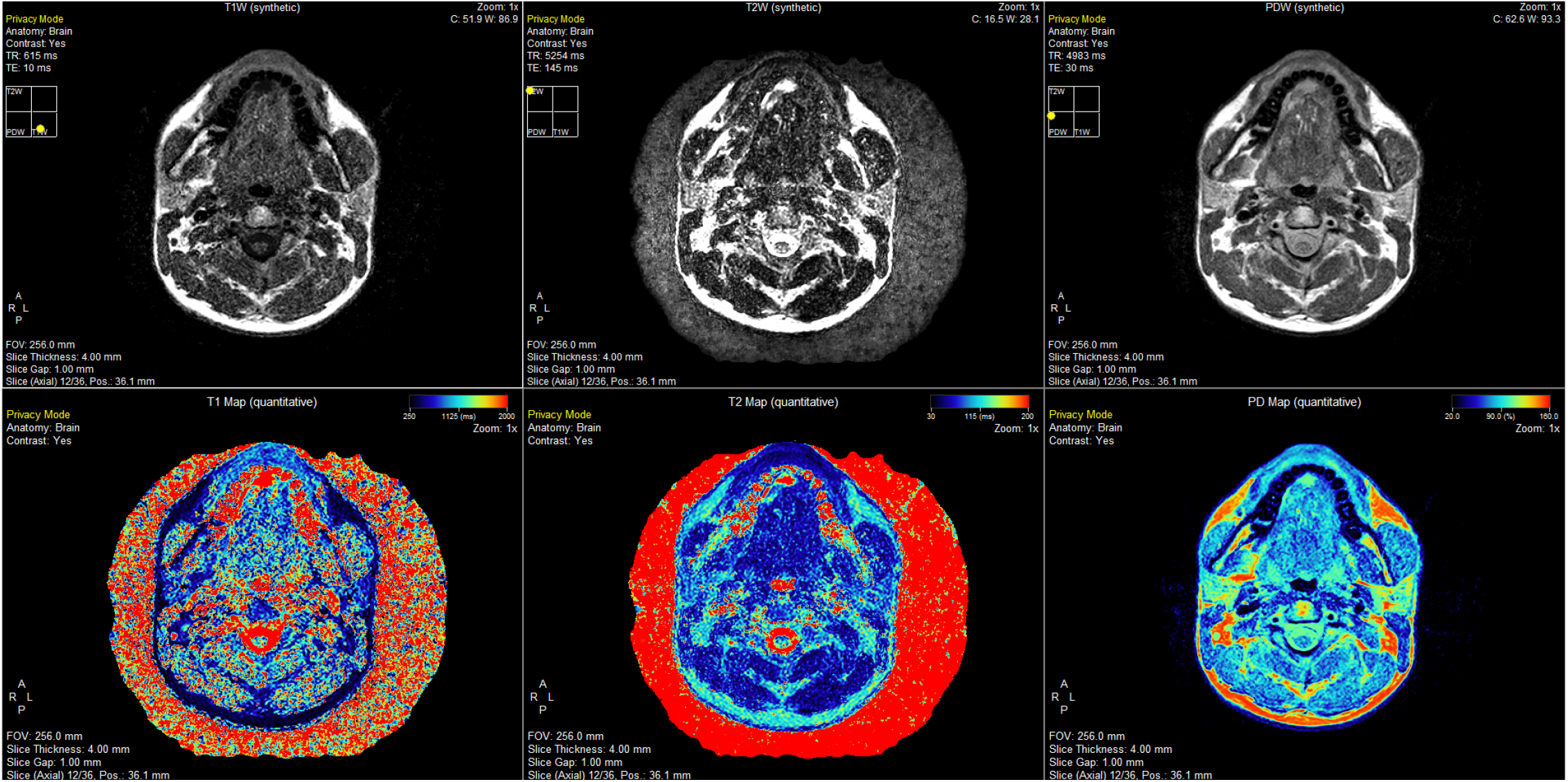
All quantitative (synthetic) and quantitative contrast maps, including T1-/T2-/PD-Weighted and quantitative T1/T2/PD derived from SyntheticMR^32^.

#### c. Dynamic Contrast Enhanced (DCE)

Physiologically, DCE can give a quantitative estimation of the perfusion between the blood vessel plasma and the surrounding extravascular extracellular space (EES) as both a volume (K^trans^) and rate (k_ep_), as well as the volumes of blood (k_b_), blood plasma (v_p_), and EES (v_e_). Additionally, derived parameters such as the amount of flow and permeability in physiological units can also be investigated using DCE. However, one complication of DCE is its un-standardized and technically complicated reconstruction methodology leading to problems of reproducibility across research groups and dependency on processing tool^41^.

Besides being used for direct diagnosis of head and neck cancers, DCE has been used to predict regional failures in head and neck cancer patients with chemoradiation therapy^42^ and identify the features and physiological explanations for these failures^43^. Other clinical applications in head and neck cancers include prediction of responders to chemoradiation therapy^44^ and sequalae in normal tissues such as those in the mandible for osteoradionecrosis^45^.

**Figure 5.**
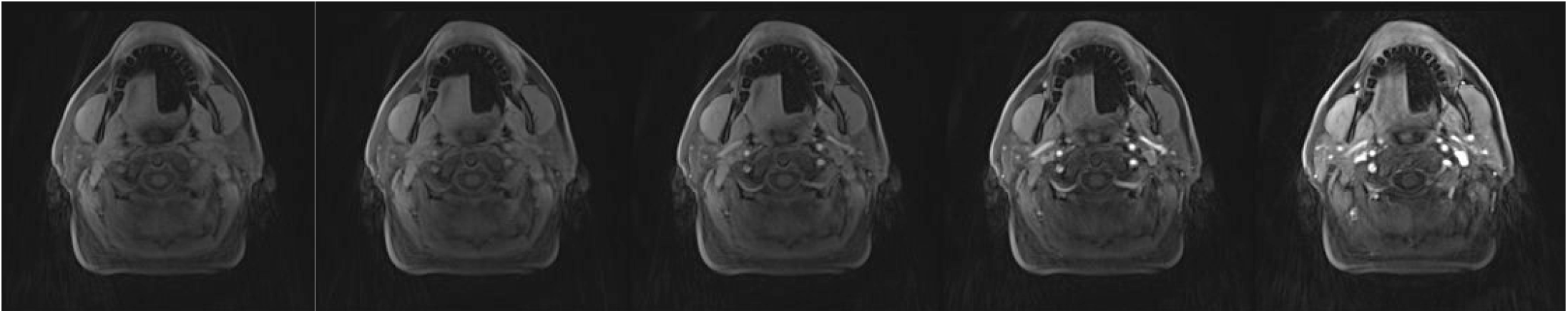
Panel showing dynamic enhancement time series (left to right).

## Discussion

When performing an MRI protocol optimization, it is essential to involve all parties which may interact with updated protocol to understand all its possible weaknesses. For example, in our updated protocol described here, the physician wanted coverage from the top of the head to just below the shoulders, an increase in coverage from the previous protocol. Without directly consulting the MRI technologists, we would have continued to scan with the increased coverage and seen significant SNR drop-off around the shoulders; however, they suggested that we add another anterior coil to cover the shoulders to prevent this issue.

Additional techniques not covered in our MR simulation protocol provide promise in becoming a piece of future iterations, specifically for the head and neck. The first is MR spectroscopy, which has been utilized in head and neck cancers to detect higher levels of amino acids in tumor^46^ and to predict response to chemotherapy and radiation therapy^47^. However, their widespread use clinically has yet to be fully realized due to their lack of reproducibility and technical challenges to achieve high uniforming in the head and neck region while maintaining feasible acquisition times, especially for multi-voxel / chemical shift imaging applications. Secondly, coherent gradient echo with balanced refocusing, also known as steady-state imaging (i.e., “TrueFISP” on Siemens, “FIESTA” on GE, and “b-FFE” on Philips), has been employed for trigeminal nerve visualization^48^.

Finally, MR lymphangiography is a promising technique to visualize the lymphatic system within a patient which can help to characterize lymph drainage pathways and lymph node localization and response^49^. This can be combined with ferumoxytol to provide further enhancement for nodal involvement in cancers^50^.

This work has several limitations. Although a rigorous statistical analysis for the quantitative accuracy, repeatability, and reproducibility of the functional / quantitative sequences were not provided in this study, the MDME sequence has been evaluated in previous study^4^ and the BLADE DWI sequence will be evaluated in a future study in comparison to our previous EPI-based technique. It is important to note that while GE’s MAGiC or Siemens’ SyMRI are highly successful in brain imaging, their performance drops significantly when moving into skull base and head and neck regions. MDME is prone to susceptibility artifact, involuntary motion from carotid pulsation, breathing, or swallowing, and irregular geometry of neck, causing magnetic field non-uniformity. Additionally, almost all sequences were acquired axially, while the T1-weighted UTE was acquired in the sagittal plane. Thus, multiplanar reconstruction is required for clinical reads in radiology. Future work will increase feasibility assessment in both healthy volunteers and head and neck cancer patients.

## Data Availability

All data produced in the present work are contained in the manuscript.

## CRediT Author Statement

Conceptualization: NAW, LM, JW, CDF. Methodology: NAW, LM, JTC, VTD, JW, JP, AL, CDF. Software: NAW, LM. Validation: NAW, LM, YD, JW, JP, AL, CDF. Formal analysis: NAW, LM. Investigation: NAW, LM, JTC, VTD. Resources: JTC, VTD, KS, KL, MC, JP, AL, AG, DR, CDF. Data Curation: NAW, LM. Writing – Original Draft: NAW, LM. Writing – Reviw & Editing: NAW, LM, JTC, VTD, YD, JW, KS, KL, MC, JP, AL, AG, DR, CDF. Visualization: NAW, LM. Supervision: CDF. Project Administration: CDF. Funding Acquisition: NAW, LM, CDF.

